# Effects of myopia and glaucoma in the prelaminar neural canal and anterior lamina cribrosa using 1060-nm swept-source optical coherence tomography

**DOI:** 10.1101/2021.09.20.21263733

**Authors:** Sieun Lee, Morgan Heisler, Dhanashree Ratra, Vineet Ratra, Paul J Mackenzie, Marinko V Sarunic, Mirza Faisal Beg

## Abstract

**Purpose:** Investigate the effects of myopia and glaucoma in the prelaminar neural canal and anterior lamina cribrosa using 1060-nm swept-source optical coherence tomography

**Design:** Retrospective, cross-sectional study

**Methods:** *Setting:* Clinical practice

*Patient or study population:* 19 controls (38 eyes); 38 glaucomatous subjects (63 eyes). Inclusion criteria for glaucomatous subjects: i) optic disc neural rim loss; ii) peripapillary nerve fibre layer (NFL) loss on spectral domain optical coherence tomography (SD-OCT); iii) glaucomatous visual field defect with an abnormal pattern standard deviation (P<.05); iv) stable SD-OCT, visual field, and optic disc clinical examination for 6 or more months. Inclusion criteria for control subjects: no evidence of retinal or optic nerve pathology. Exclusion criteria: i) retinal diseases or optic neuropathy other than primary open-angle glaucoma; ii) intraocular pressure less than 10 mmHg or higher than 20 mmHg; iii) ocular media opacities; iv) any surgery-related complication deemed inappropriate for the study.

*Intervention or observation procedures:* Swept-source optical coherence tomography

*Main Outcome Measure(s):* Bruchs membrane opening (BMO) and anterior laminar insertion (ALI) dimension, prelaminar neural canal dimension, anterior lamina cribrosa surface (ALCS) depth

*Results:* Glaucomatous eyes had more bowed and nasally rotated BMO and ALI, more horizontally skewed prelaminar neural canal, and deeper ALCS than the control eyes. Increased axial length was associated with a wider, longer, and more horizontally skewed neural canal, and decrease in the ALCS depth and curvature.

*Conclusion:* Our findings suggest that glaucomatous posterior bowing or cupping of lamina cribrosa can be significantly confounded by the myopic expansion of the neural canal. This may be related to higher glaucoma risk associated with myopia from decreased compliance and increased susceptibility to IOP-related damage of LC being pulled taut.

## Introduction

Glaucoma is a leading cause of blindness in the world and a multi-factorial group of diseases involving progressive damage to the retinal ganglion cells (RGCs) and axons resulting in irrevocable vision loss. Although the etiology and mechanism of glaucoma are not yet fully understood, elevated intraocular pressure (IOP) is an important risk factor and treatment target. In experimental and clinical studies,^1–3^ lamina cribrosa (LC) in the optic nerve head (ONH) has been shown as the main site of RGC axonal damage in early glaucoma, and the IOP-related deformation, remodeling, and mechanical failure of the ONH connective tissues are proposed as the defining pathophysiology of the disease.^4–6^ These alterations include posterior laminar deformation,^7,8^ scleral canal expansion,^8,9^ posterior migration of anterior laminar insertion (ALI),^10,11^ laminar thickness change,^7,12^ and posterior bowing of the peripapillary sclera.^2^ The biomechanical paradigm suggests that the response of an individual ONH to a given level of IOP depends on its structural factors, affecting the disease susceptibility, occurrence, and progression.

In myopia, the eye elongates in the anterior – posterior (axial) direction, causing poor focus in distance vision. The myopic elongation, particularly in high myopia, affects the ONH structure.^13–17^ In the standard glaucoma examination of the ONH and peripapillary region using ophthalmoscopy and optical coherence tomography (OCT), myopia can confound the assessment with shallow cupping, pallor in the neuroretinal rim, and retinal nerve fiber layer thinning.^18–20^ Furthermore, high myopia has been associated with increased susceptibility to glaucoma in several population-based studies.^21–23^ These findings suggest that myopic deformation of the ONH structure may affect the biomechanism of glaucoma development and progression.

OCT has now become a standard technology in ophthalmic clinics, providing high resolution and cross-sectional 3D images of the retina. Compared to a conventional spectral domain OCT, a swept-source OCT (SS-OCT) with a 1060-nm wavelength source allows for visualization of deeper structures in the ONH. Using SS-OCT, we have previously published on the morphological characteristics of the ONH and peripapillary region in myopic normal and glaucomatous subjects.^24–26^ We showed that axial length was a significant factor in the shape of Bruch’s membrane opening (BMO) and the degree of posterior bowing of the peripapillary BM. In the present study, we extend the investigation to the prelaminar neural canal between BMO and ALI and anterior lamina cribrosa morphology in healthy and glaucomatous myopic subjects to further examine the structural effects of myopia and glaucoma in the ONH.

## Materials and Methods

### Participants

The study was conducted in accordance with the Declaration of Helsinki and approved by the institutional research ethics boards of Simon Fraser University (SFU) and University of British Columbia (UBC). Myopic participants with and without glaucoma were included following a full clinical examination by a fellowship-trained glaucoma specialist (PJM), including dilated stereoscopic imaging, IOP measurement using Goldmann applanation tonometer, and reproducible Humphrey SITA-Standard white-on-white visual field. Axial length was measured using Zeiss IOLMaster (Carl Zeiss Meditec, Jena, Germany).

The glaucomatous eyes included in the study met the following criteria: i) evidence of optic disc neural rim loss on clinical examination; ii) evidence of peripapillary NFL loss on spectral domain optical coherence tomography (SD-OCT); iii) glaucomatous visual field defect with an abnormal pattern standard deviation (P<.05); iv) stable SD-OCT, visual field, and optic disc clinical examination for 6 or more months. The nonglaucomatous eyes included in the study showed no evidence of retinal or optic nerve pathology. From both groups, eyes were excluded based on following criteria: i) retinal diseases or optic neuropathy other than primary open-angle glaucoma, including myopic degeneration; ii) IOP lower than 10 mmHg or greater than 20 mmHg; iii) ocular media opacities; iv) any surgery-related complication that the investigators determined inappropriate for the study.

### Image acquisition and processing

The participants were imaged at the Eye Care Centre, Vancouver General Hospital, using a custom swept-source optical coherence tomography (SS-OCT) system with a 1060-nm light source, built by Biomedical Optics Research Group at SFU. The system had 100 KHz A-scan rate and 1.6 second imaging time with improved visualization of deeper ONH structures compared to conventional 800-nm range commercial OCT systems. The acquired volumetric images consisted of 400 B-scans with voxel dimension of 1024 × 400. The axial resolution was 2.8 µm and the lateral resolution was 11 to 20 µm, depending on the axial length of the eye. Axial motion correction and bounded variance smoothing was applied to improve the image quality.

### Segmentation

Bruch’s membrane (BM), Bruch’s membrane opening (BMO), anterior laminar insertion (ALI), and anterior lamina cribrosa surface (ALCS) were segmented (Figure 1-A). BM was segmented automatically using a 3D graph-cut based method^25^ and the result was examined and corrected for segmentation errors by trained raters. BMO, ALI, and ALCS were segmented manually using Amira (Thermo Fisher Scientific, MA) on 80 radial slices extracted from the OCT volume, intersecting at the approximate centre of BMO and equidistanced at 2.25°.

**Figure 1.**
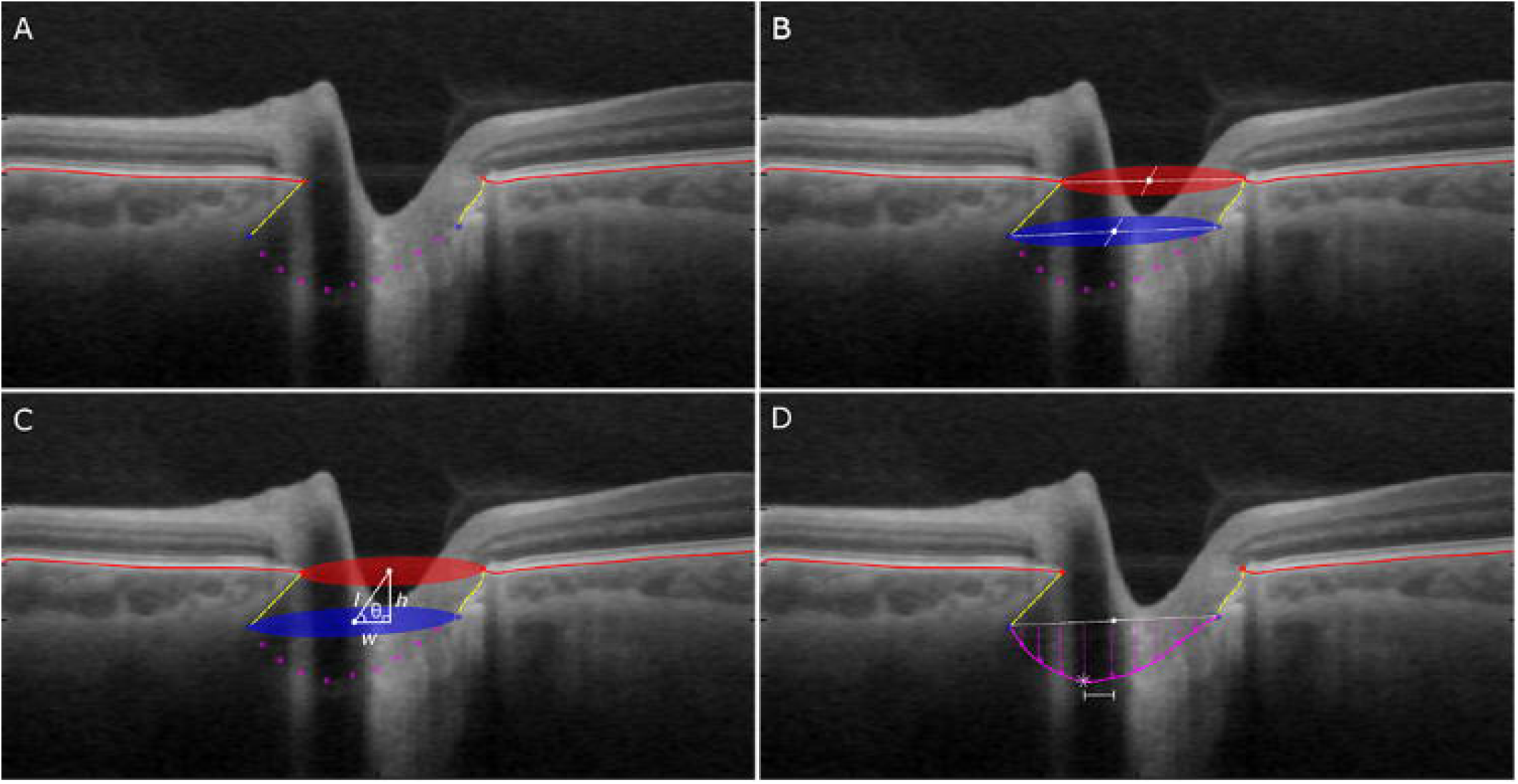
Optic nerve head shape parameterization. A) Bruch’s membrane (red curve), Bruch’s membrane opening (BMO, red dot), prelaminar neural canal (yellow curve), anterior laminar insertion points (ALIP, blue dots), and anterior lamina cribrosa surface (ALCS, pink dot). B) BMO and ALIP centroids (white dot) and major and minor axes (white dotted line). C) PNC height (h), width (w), length (l), and angle (θ). D) ALCS depth (magenta arrows), ALIP centroid (white dot), ALCS deepest point (white asterisk), and off-centre distance (white line).

### Shape Parameterization

The prelaminar region was approximated as an oblique cylinder, open at the top by BMO, and closed at the bottom by ALCS outlined by ALI. Parameters were defined to characterize the geometry of the cylinder and grouped into BMO and ALI parameters, prelaminar neural canal parameters, and ALCS parameters. BM plane was used as the reference, computed by principal component analysis (PCA) on BM points at 2 mm away from BMO.

#### BMO and ALI

BMO and ALI were modelled as ellipses and measured for their sizes, lateral elongation, axial nonplanarity, and distance from BM (Figure 1-B). For geometrical analysis of BMO, an ellipse was fitted to the 160 segmented BMO points by computing the best-fit plane using PCA and fitting an ellipse to the projection of the points on the plane using the minimum squared error criterion. The area, location of the centroid, and major and minor axes were obtained from the BMO ellipse. BMO lateral elongation was characterized by eccentricity (ratio of the major and minor axes) and direction in the enface plane (angle between the BMO ellipse major axis and superior-inferior axis). BMO nonplanarity was calculated as the mean normal distance of the segmented BMO points to its own best-fit plane. This was also normalized to relative nonplanarity by dividing BMO nonplanarity by BMO area. BMO distance from BM was measured as the normal distance between the BM plane and BMO centroid. Same parameters were computed for ALI. The ellipse fitting approach mitigates the effects of heavy blood vessel shadowing in the ONH region and resulting difficulty of segmenting BMO or ALI points in some slices.

#### Prelaminar Neural Canal

Prelaminar neural canal was measured for the dimension (length, height, width) and direction (axial and lateral angles), and the difference between the anterior and posterior ends (expansion, bowing, twist) as follows (Figure 1-C). First, canal axis was defined as the vector between the BMO centroid and ALIP centroid. Canal length was defined as the length of the vector, and the height and width as the lengths of the axial and lateral components of the vector. Canal angle was measured as the angle between the canal axis and the vertical (axial) axis of the image, and the enface canal direction was measured as the angle between the lateral component of the canal axis and the superior-inferior axis. The differences between the BMO and ALIP areas (posterior expansion), eccentricities, nonplanarity, and major axes (twist) were also measured.

#### ALCS

ALCS was measured for the degree of surface cupping (depth, curvature), and off-centeredness of the deepest point (Figure 1-D). The parameters were measured for the manual ALCS segmentation points and their least square-fit quadratic polynomial surface.

Mean ALCS depths were computed as the average normal distances from the ALI plane to the segmented points and to the fitted surface on a 0.01 mm grid. The mean depth was also normalized by dividing by the ALI area. For the fitted surface, mean curvature and the relative enface position of the ALCS minimum (defined as the deepest point on the surface) to the ALI centroid were measured.

### Statistical Analysis

The nonglaucomatous and glaucomatous groups were compared using generalized estimation equation analysis, accounting for the inter-eye correlation and including age and axial length in the model to adjust for any confounding. The effects of age and axial length in the nonglaucomatous group and the effects of age, axial length, and visual field mean deviation (MD) in the glaucomatous group on the shape parameters were evaluated using a mixed effect model with a random subject effect for the inter-eye correlation. SPSS 24 (IBM Corp. Released 2016. IBM, Armonk, NY) and SAS (SAS Institute Inc., Cary, NC) were used for the analysis.

## Results

The characteristics of the study population are listed in Table 1. There was a significant difference in the age and visual field mean deviation (MD) between the nonglaucomatous and glaucomatous groups.

**Table 1.** Participant summary. Number of subject, Number of Eyes, and mean ± SD (minimum − maximum) Age, Axial Length,and visual field index (Mean Deviation) of nonglaucomatous and glaucomatous participants

|  | <u>Nonglaucomatous</u> | **Glaucomatous** | **P** |
| --- | --- | --- | --- |
| **Number of Subjects** | 19 (10 females) | 38 (15 females) |  |
| **Number of Eyes** | 38 | 63 |  |
| **Age** | 37.3 ± 13.9 (20-61) | 61.2 ± 12.5 (30-84) | <.001 |
| **Axial Length (mm)** | 25.1 ± 1.42 (23.1 – 28.42) | 25.9 ± 1.78 (22.5-31.28) | - |
| **Mean Deviation (dB)** | -0.91 ± 1.00 (-.06 – 0.97) | -11.9 ± 8.82 (-33.6 – 0.83) | <.001 |

The mean results for Bruch’s membrane opening (BMO), anterior laminar insertion (ALI), prelaminar neural canal (PNC) and anterior lamina cribrosa surface (ALCS) for nonglaucomatous and glaucomatous eyes are shown in Table 2.

**Table 2.**
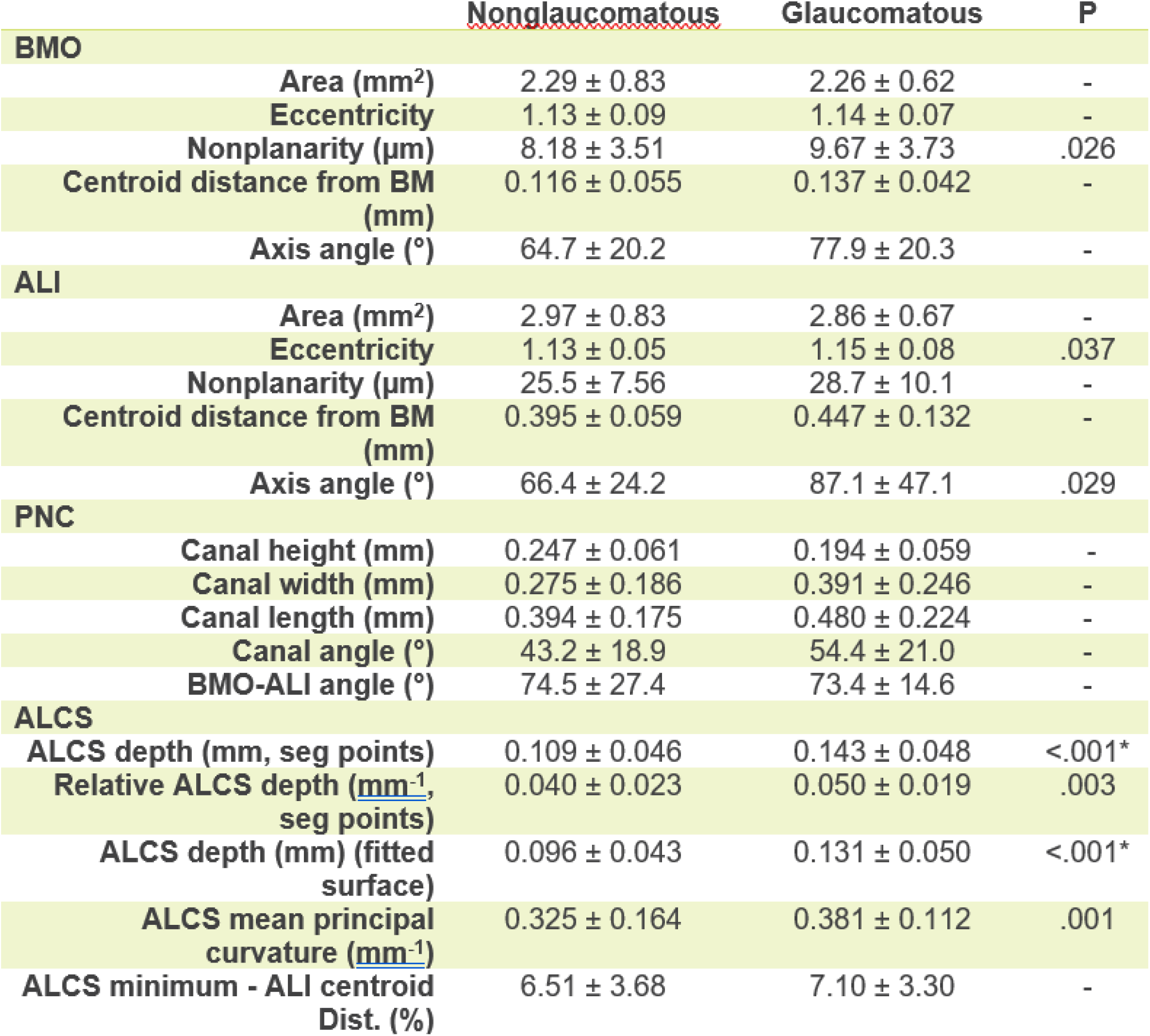
Mean measurements of Bruch’s membrane opening (BMO), anterior laminar insertion point (ALI) ellipse, prelaminar neural canal (PNC) and anterior laminar surface (ALCS) for healthy and glaucomatous eyes. An asterisk indicates p < .001.

Significant p-values (< .05) from the general linear models of the measurements with age and axial length (AL) as predictors for nonglaucomatous eyes and with age, AL, and visual field index (MD) are shown in Table 3.

**Table 3.** Significant p-values from general linear models of Bruch’s membrane opening (BMO), anterior laminar insertion (ALI), prelaminar neural canal (PNC), and anterior laminar surface (ALCS) measurements with age, axial length (AL), and visual field index (MD) as predictors of nonglaucomatous and glaucomatous eyes. Red indicates the predictor was negatively associated with the variable.

|  | <u>Nonglaucomatous</u> |  | Glaucomatous |  |  |
| --- | --- | --- | --- | --- | --- |
|  | Age | AL | Age | AL | MD |
| **BMO** |  |  |  |  |  |
| Area (mm<sup>2</sup>) | - | .008* | - | .003* | - |
| Eccentricity | - | .010 | - | .018 | - |
| Nonplanarity (μm) | - | - | - | - | .039 |
| Centroid distance from BM (mm) | - | .018 | - | - | .004 |
| Axis angle (°) | - | .009* | - | - | - |
| **ALI** |  |  |  |  |  |
| Area (mm<sup>2</sup>) | - | .014 | - | .011 | - |
| Eccentricity | - | - | - | - | - |
| Nonplanarity (μm) | - | - | - | - | - |
| Centroid distance from BM (mm) | - | - | - | .030 | <.001* |
| Axis angle (°) | - | - | - | - | - |
| **PNC** |  |  |  |  |  |
| Canal height (mm) | - | .004* | - | - | - |
| Canal width (mm) | - | <.001* | - | .025 | - |
| Canal length (mm) | - | <.001* | - | - | .025 |
| Canal angle (°) | - | <.001* | - | - | - |
| BMO-ALI angle (°) | - | - | - | - | .036 |
| **ALCS** |  |  |  |  |  |
| ALCS depth (mm, points) | - | .037 | - | - | .002* |
| Relative ALCS depth (mm<sup>-1</sup>, points) | - | .011 | - | .005* | <.001* |
| ALCS depth (mm) (fitted surface) | - | .047 | - | - | - |
| ALCS mean principal curvature (mm<sup>-1</sup>) | - | .002* | - | - | - |
| ALCS minimum - ALI centroid Dist. (%) | - | - | - | - | - |

### Bruch’s Membrane Opening and Anterior Laminar Insertion

BMO **area** and **eccentricity** were not significantly different between the nonglaucomatous and glaucomatous eyes (Table 2) but longer axial length was associated with a larger and more elliptical BMO was associated in both groups (Table 3). Compared to BMO, ALI area was larger and similarly associated with longer axial length in both nonglaucomatous and glaucomatous eyes (Table 3).

BMO and ALI **nonplanarity** is visualized for a representative eye in Figure 2. On the right side, the normal distance of the BMO and ALI points are plotted in reference to their own best-fit planes, starting from the temporal axis and progressing in the clockwise direction. The plot shows BMO and ALI shape in the axial direction, with nasal-temporal ends being the most anterior, and inferior-superior ends being the most posterior. The enface views on the left side of the figure show the points anterior (green) and posterior (red) to the reference planes. The enface images also demonstrate the long ends of the BMO and ALI being more anterior than the short ends. The nonplanarity in ALI was in general greater than in BMO, following a similar pattern but with some rotation in the enface axis angle. Figure 3 shows the BMO and ALI nonplanarity plotted for all eyes in nonglaucomatous and glaucomatous groups. Between the two groups, BMO nonplanarity was significantly greater in the glaucoma eyes (Table 2) and associated with glaucoma severity (Table 3).

**Figure 2.**
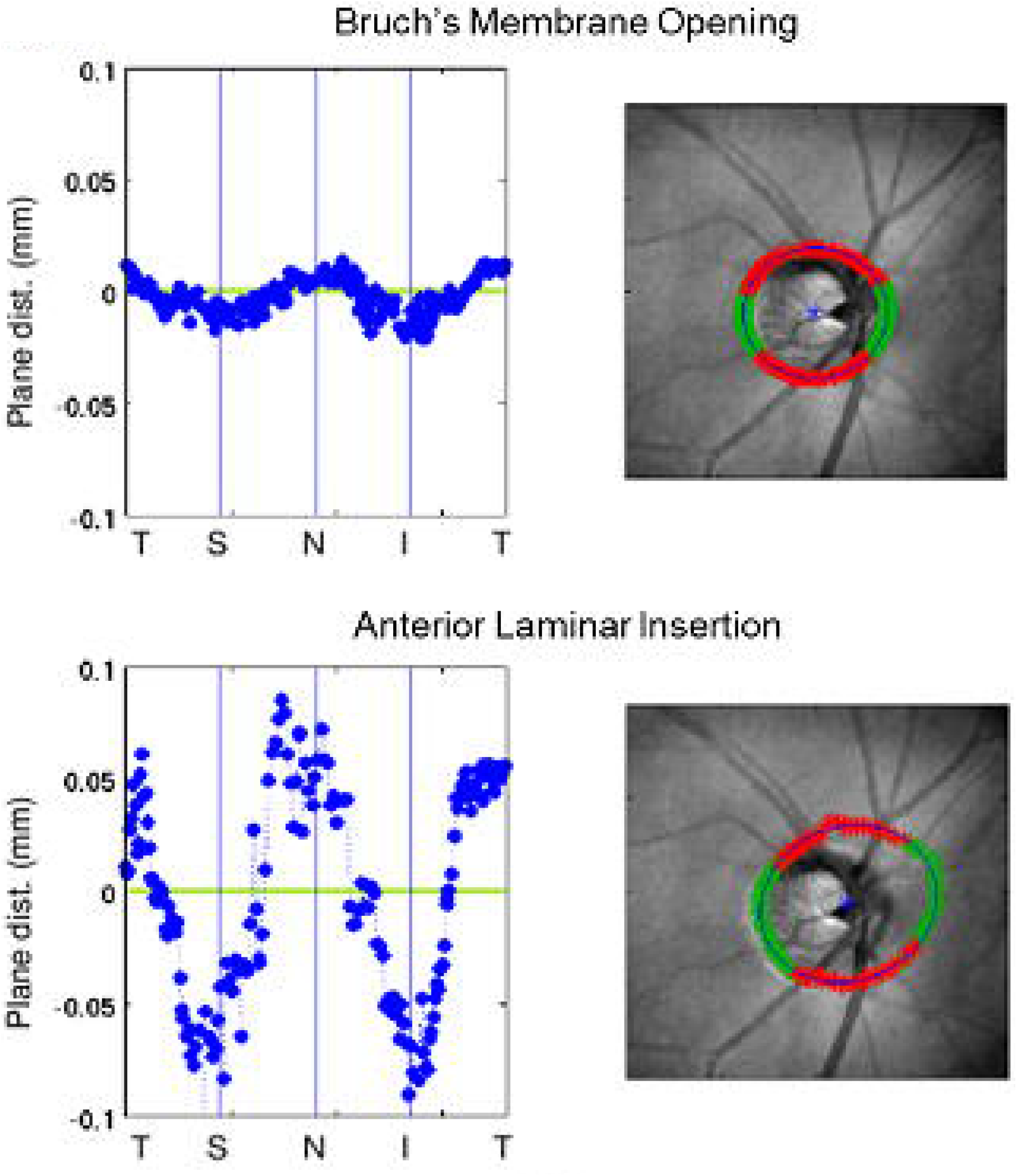
Bruch’s membrane opening (BMO) and anterior laminar insertion (ALI) nonplanarity.

**Figure 3.**
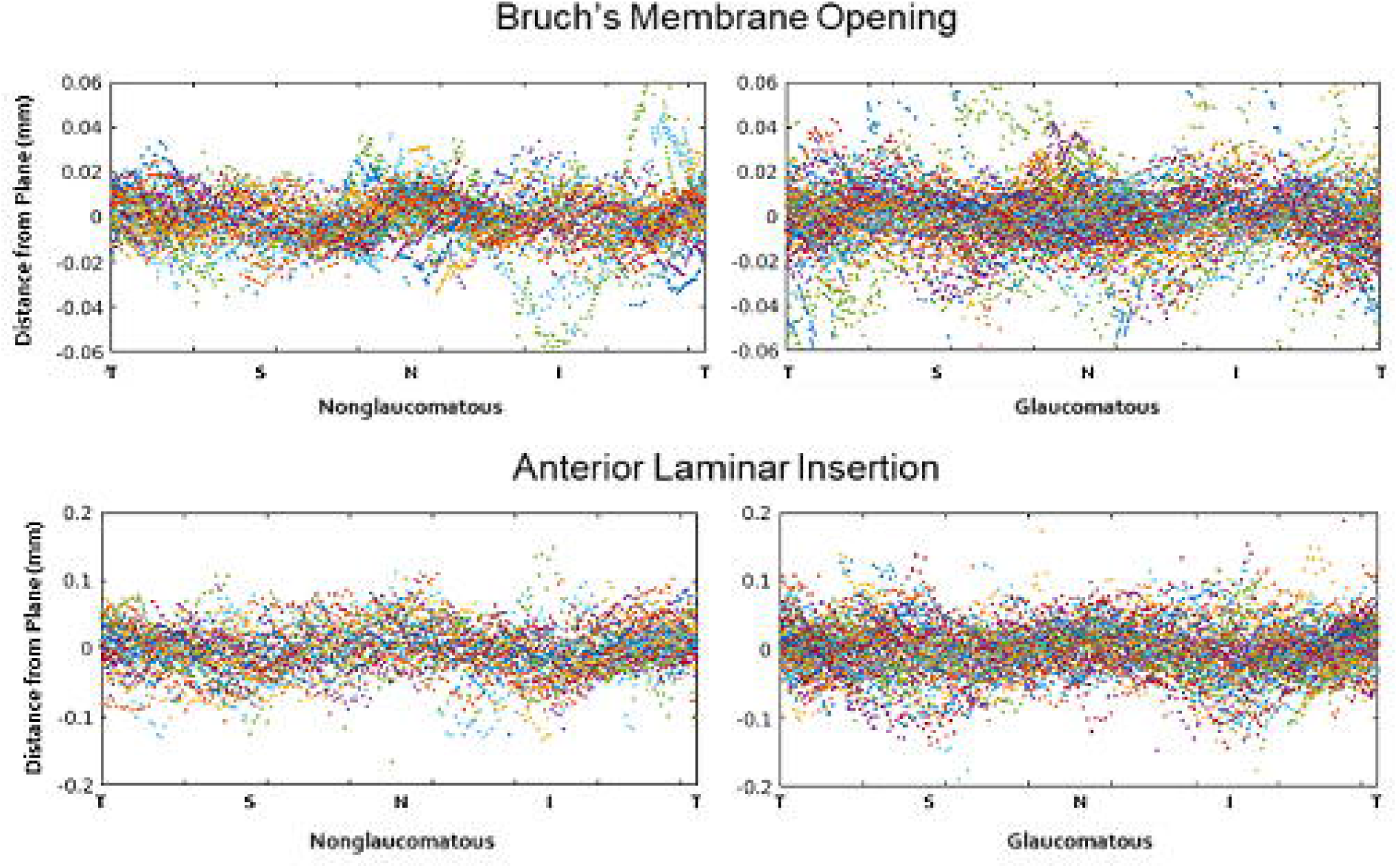
Bruch’s membrane opening points (top) and anterior laminar insertion points (bottom) in reference to the best-fit plane for nonglaucomatous and glaucomatous eyes.

The **distance of the BMO centroid from BM plane** in the posterior direction, which can be contributed by both deformation and displacement of BMO, was negatively associated with axial length in nonglaucomatous eyes and positively associated with glaucoma severity in glaucomatous eyes (Table 3). The distance of the ALI centroid from BM plane was significantly greater in the glaucomatous eyes (Table 2), and positively associated with both axial length and glaucoma severity (Table 3).

Table 4 shows the frequency of **BMO axis direction**, grouped in six sectors with equal angles with reference to superior (S), inferior (I), nasal (N), and temporal (T) orientations. In both groups, BMO axis most frequently lied in the nasal-superior-nasal to nasal sectors. This directionality corresponds with the BMO bowing, and shows that BMO tends to be the most anterior on its longest ends (NSN-TIT to N-T) and most posterior on its shortest ends (IIN-SST to S-I). More BMO axes were in the nasal-temporal sectors for the glaucomatous eyes. BMO axis direction was associated with increased axial length (Table 3), suggesting BMO axis rotation is influenced by both myopia and glaucoma, and BMO nonplanarity and axis rotation may be related in glaucoma.

**Table 4.** Frequency of Bruch’s membrane opening (BMO) axis directions.

|  | **S-I** | **SSN-IIT** | **NSN-TIT** | **N-T** | **NIN-STS** | **IIN-SST** |
| --- | --- | --- | --- | --- | --- | --- |
| **Nonglaucomatous (n = 38)** | 2 | 8 | 16 | 10 | 1 | 1 |
| **Glaucomatous (n = 63)** | 2 | 8 | 18 | 26 | 6 | 3 |

Table 5 shows the frequency of ALI axis direction. As in BMO, the ALI axis also mainly lied in the nasal-superior-nasal to nasal sectors, corresponding to ALI nonplanarity pattern. There was overall greater variability in the ALI axis of glaucomatous eyes and larger mean axis angle (Table 2).

**Table 5.**
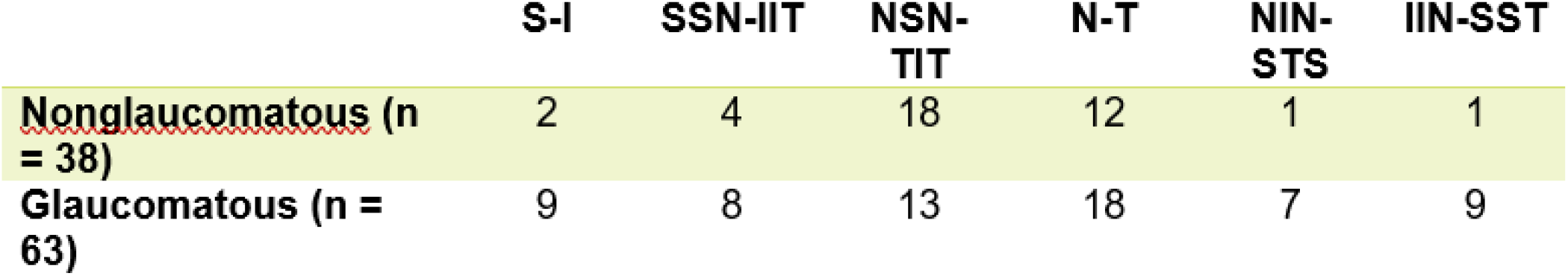
Frequency of anterior laminar insertion (ALI) axis direction

|  | **S-I** | **SSN-IIT** | **NSN-TIT** | **N-T** | **NIN-STS** | **IIN-SST** |
| --- | --- | --- | --- | --- | --- | --- |
| **Nonglaucomatous (n = 38)** | 2 | 4 | 18 | 12 | 1 | 1 |
| **Glaucomatous (n = 63)** | 9 | 8 | 13 | 18 | 7 | 9 |

### Prelaminar Neural Canal

In the nonglaucomatous eyes, prelaminar neural canal (PNC) was longer and more skewed with longer axial length. In the glaucomatous eyes, longer axial length was associated with more skewed canal, and greater glaucoma severity was associated with longer canal, likely due to the posterior cupping and displacement of ALI. Glaucoma severity was also associated with decrease in the BMO-ALIP axis angle difference (Table 3), possibly reflecting the apparent glaucomatous rotation of BMO axis angle toward nasal-temporal axis (Table 4). Table 6 shows the frequency of BMO-to-ALI direction.

**Table 6.**
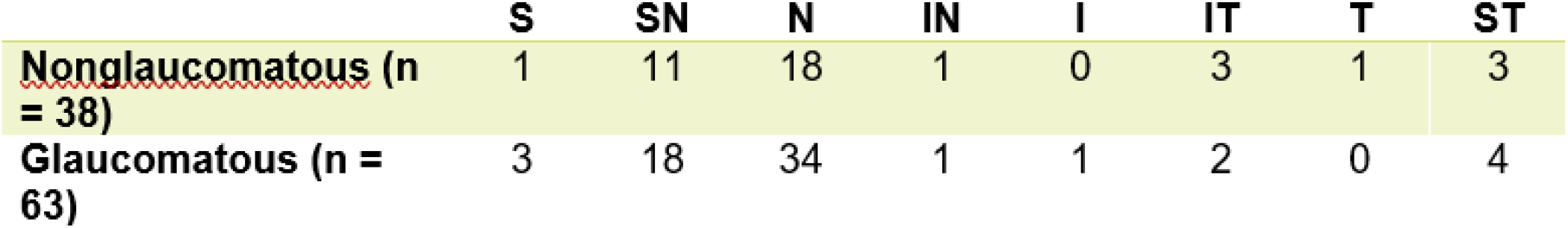
Frequency of BMO to ALI direction

### Anterior Laminar Surface

Anterior laminar surface (ALCS) was significantly deeper in the glaucomatous eyes (Table 2) and ALCS depth positively correlated with glaucoma severity (Table 3).

However, in both nonglaucomatous and glaucomatous eyes, axial length was negatively correlated with ALCS depth. In nonglaucomatous eyes, longer axial length was significantly associated with less ALCS curvature of the fitted surface.

Table 7 shows the frequency of the location of the ALCS surface minimum. In both groups, ALCS minimum was mainly located in the superior, superior-nasal, and nasal sectors, corresponding to the directions of BMO and ALI elongation and the prelaminar neural canal.

**Table 7.** Frequency of anterior laminar surface minimum location

|  | **S** | **SN** | **N** | **IN** | **I** | **IT** | **T** | **ST** |
| --- | --- | --- | --- | --- | --- | --- | --- | --- |
| **Nonglaucomatous (n = 38)** | 7 | 15 | 7 | 1 | 2 | 3 | 4 | 2 |
| **Glaucomatous (n = 63)** | 17 | 27 | 8 | 0 | 2 | 0 | 4 | 8 |

## Discussion

Development of myopia typically begins between age six and fourteen, and the progression stops or slows at the end of adolescence as the general physical growth ends.^27,28^ The axial elongation of the eye during this process affects the structure of the optic nerve head (ONH). On the other hand, most common adult glaucoma occurs in mid-to-late adulthood, with intraocular pressure (IOP)-related deformation and remodeling of the ONH playing a key role in the pathophysiology. Myopia has been associated with increased susceptibility to glaucoma in several population studies,^21–23^ and the relationship between myopia and glaucoma involves the myopic ONH structure, the response of the myopic ONH to aging and IOP elevation, and how these affect the onset and progression of glaucoma.

### Literature on myopia and optic nerve head

- **Optic disk** Early histological studies found that optic disks were **larger** and **more oval** in highly myopic eyes.^29,30^ Recent *in vivo* studies showed increasing axial length was associated with **oblique or skewed insertion of the optic nerve into the globe**^31^ in fundus photography ^32,33^ and in 3D optical coherence tomography (OCT) ^17,34,35^. The resulting rotation of the major axis of the optic disk in the enface plane (**optic disk torsion**) has been associated with **location of visual field defects** in glaucoma patients^33,36^. A caveat is that the usage of the terms such as optic disk *tilt, ovality, torsion*, and *skew* have varied across the literature, especially between studies based on 2D fundus photographs and those based on 3D OCT, and the correlation between the 2D and 3D measurements for the supposedly same anatomical measurements has been shown to be limited.^35,37^
- **Lamina cribrosa and sclera** Lamina cribrosa (LC) was found to be significantly **thinner in highly myopic eyes** than in non-highly myopic eyes in both glaucoma and control groups, resulting in a shorter distance between the intraocular space and cerebrospinal fluid space and increased translaminar pressure gradient.^12^ The same experiment also showed that among highly myopic eyes, **presence of glaucoma was associated with thinner LC** than in non-highly myopic eyes. Biomedical models of the sclera further associated myopia with **scleral thinning**,^38,39^ **tissue loss**,^39^ **failure at a lower load**,^38^ **and increase in small diameter collagen fibrils**.^39^

### Literature on myopia and glaucoma

- Among glaucoma patients, high myopia was identified as a significant risk factor for subsequent **visual field loss**,^40,41^ and in examination of glaucomatous optic nerve fiber loss using color stereo optic disc photographs, **optic nerve damage** was shown to be more significant in highly myopic eyes than in non-highly myopic eyes.^42^
- **RNFL thickness** OCT-based measurement of peripapillary retinal nerve fiber layer (RNFL) thickness, a key measurement of glaucoma severity, was also shown to be thinner in healthy myopic subjects.^19,43,44^ However, because of the myopic expansion of the optic disk and Bruch’s membrane opening, RNFL thickness measurements based on the distance from the centre of the optic disk may be located closer to the BMO for myopic subjects than non-myopic subjects.

### Our study

**Summary** In this paper, we presented a comprehensive 3D shape characterization and analysis of the anterior laminar region in myopic eyes with and without glaucoma using images from a custom swept-source optical coherence tomography (SS-OCT) system with a 1060-nm light source. Bruch’s membrane opening (BMO), anterior laminar insertion (ALI), prelaminar neural canal (PNC), and anterior lamina cribrosa surface (ALCS) were segmented from the images and analyzed for their relationships with age, axial length, and glaucoma severity. The age-adjusted results showed that the glaucomatous eyes had more bowed and nasally rotated BMO and ALI, more horizontally skewed PNC, and deeper ALCS than the nonglaucomatous eyes. General linear model analysis showed that increased axial length was a significant factor across the anterior laminar region, most notably with a wider, longer, and more horizontally skewed neural canal, and decrease in the ALCS depth and curvature. Such 3D evaluation of the ONH enhances the understanding of the peri-laminar structures and anatomy and can be a complementary tool to the conventional methods of assessment of glaucomatous or suspected glaucomatous eyes.

Serial SD-OCT scans in experimental glaucoma showed that changes in the deeper structures such as the laminar and prelaminar tissues precede other obvious glaucomatous changes such as thinning of the RNFL.^45^ The age-adjusted results showed that the glaucomatous eyes had more bowed and nasally rotated BMO and ALI, more horizontally skewed PNC, and deeper ALCS than the nonglaucomatous eyes, which may be a result of the increased intraocular pressure. On the anatomical parameterization, the authors noted that the neural canal opening plane was a valid reference point for assessing and quantifying the ONH parameters, and although the whole neural canal along with the LC may shift posteriorly due to glaucoma, the centroid of the neural canal opening remains stable. Sigler et al.^46^ found that prelaminar canal depth, when measured from the BMO to ALCS, was positively correlated with peripapillary choroidal thickness and this should be considered in modeling. Our study did not explicitly look at the extent of contribution of the peripapillary choroidal thickness to the neural canal length of depth. However, studies have demonstrated choroidal thinning in myopia^47^ and this would mean that the neural canal lengthening we observed in the myopic eyes were in spite of any associated choroidal thinning. We also note that our ALCS depth and curvature were defined using only the anterior laminar insertion points and anterior lamina cribrosa surface, without referencing Bruch’s membrane or BMO, in order to more accurately capture the shape of the ALCS.

**Myopia & glaucoma’s effects in the ONH structure - Compare & Contrast** Our study showed that myopia and glaucoma both affect the ONH structure; however, we note that their underlying mechanisms are fundamentally different. In myopia, the enlarged BMO and ALI areas suggests that the myopic axial elongation of the eye expands the optic neural canal, with the force acting mainly along the scleral wall. In glaucoma, the intraocular pressure acting on the ONH has some laminar component but compared to the myopic expansion it acts more in the axial direction. This simplified model can be useful for interpreting the current result of the ALCS depth being negatively associated with axial length but positively associated with glaucoma severity. In myopia, the enlarging of the canal pulls the LC taut, decreasing its curvature. This model is in line with previous studies in which myopic LC and sclera were shown to be thinner than in nonmyopic eyes.^12,38,39^ In glaucoma, the biomechanical model includes posterior bowing or “caving in” of the LC, and in our study the glaucomatous eyes indeed had deeper ALCS. Increased visual field deviation was also associated with deeper ALCS.

Another parameter affected by both myopia and glaucoma was the enface angle of the BMO and ALI axes, which were more horizontal (closer to temporal-nasal axis) with both greater axial length and glaucoma severity. A myopic mechanism of the phenomenon may be that the myopic expansion of the prelaminar region and in extension scleral opening occurs at a greater degree in the temporal-nasal direction than the superior-inferior direction. A glaucomatous mechanism of the same phenomenon may be that the intraocular pressure affects LC with regional variation.

**So why are myopes more susceptible to glaucomatous damage?** An implication of our study is that the posterior bowing of the LC as a structural parameter of glaucoma severity can be significantly confounded by myopia, and that the myopic LC that is pulled taut, shallower, and thinner than non-myopic eyes may be more vulnerable to mechanical failure and axonal damage inside the LC, despite the reduced global curvature. Previously, in an idealized, analytical microstructural model of the ONH, scleral canal eccentricity, LC stiffness, LC thickness, and eye radius were found to be important determinants of the stress and strain in the LC^48^. Sawada et al. reported that an increased neural canal angle, which correlated with a longer axial length in our study and a recent study by Jeoung et al.^49^, was associated with a higher number of temporal LC defects, which in turn spatially corresponded with visual field defects in open-angle glaucoma patients with myopia^50^. The authors posited that the myopic elongation of the globe may lead to stretching of the LC in the temporal periphery, causing localized strain in the LC pores that may become torn and become larger pores that are susceptible to glaucomatous strain. Such myopic stretching in the temporal direction can also explain the BMO elongation direction we observed in the current study, in which longer axial length was associated with more elongated BMO and more nasal-temporally (horizontally) oriented BMO enface axis.

Age has been reported as a factor in LC deformation. In glaucomatous eyes with the same functional loss, older eyes presented shallower LC^51^ and less LC deformation^50^ than in younger eyes. This is likely related to the age-related decrease in the mechanical compliance of LC^52^. In our study, age was not a significant factor in the shape parameters in either the nonglaucomatous or glaucomatous groups. This may be attributed to the limited sample size and age distribution, and the effects of myopia and glaucoma being larger than that of age. However, myopic stretching of LC can result in reduction of LC compliance, similarly with aging. Considering that aging and high myopia are two key risk factors in glaucoma, we may conjecture that independent of the degree of glaucomatous global LC deformation (cupping), reduced LC compliance itself, either from aging and/or myopic stretching, may be an important causal factor in the IOP-related axonal damage in glaucoma.

This can further relate to the connection between myopia and normal-tension or low-pressure glaucoma. Studies have demonstrated that the association between myopia and glaucoma were stronger at lower IOP levels than higher IOP levels,^53^ and Sigler et al.^46^ observed wider horizontal neural canal opening in the low pressure glaucoma group compared to the normal or primary open-angle glaucoma groups. In our study, similar horizontal torsion (T-N axis) and directed expansion of BMO and ALI were observed with increased axial length. These findings suggest that myopia causes characteristic structural deformation in the optic neural canal region, and this may create a risk factor especially for a certain subtype of glaucoma related to the deformation and lower IOP range.

## Data Availability

Due to the nature of this research, participants of this study did not agree for their data to be shared publicly, so supporting data is not available.

